# Individual level prediction of emerging suicide events in the pharmacologic treatment of bipolar disorder

**DOI:** 10.1101/2021.01.13.20246603

**Authors:** Sean X. Luo, Adam Ciarleglio, Hanga Galfalvy, Michael Grunebaum, Leo Sher, J. John Mann, Maria A. Oquendo

## Abstract

**Background:** Patients with bipolar disorder have a high lifetime risk of suicide. Predicting, preventing and managing suicidal behavior are major goals in clinical practice. Changes in suicidal thoughts and behavior are common in the course of treatment of bipolar disorder.

**Methods:** Using a dataset from a randomized clinical trial of bipolar disorder treatment (N=98), we tested predictors of future suicidal behavior identified through a review of literature and applied marginal variable selection and machine learning methods. The performance of the models was assessed using the optimism-adjusted C statistic.

**Results:** Number of prior hospitalizations, number of prior suicide attempts, current employment status and Hamilton Depression Scale were identified as predictors and a simple logistic regression model was constructed. This model was compared with a model incorporating interactions with treatment group assignment, and more complex variable selection methods (LASSO and Survival Trees). The best performing models had average optimism-adjusted C-statistics of 0.67 (main effects only) and 0.69 (Survival Trees). Incorporating medication group did not improve prediction performance of the models.

**Conclusions:** These results suggest that models with a few predictors may yield a clinically meaningful way to stratify risk of emerging suicide events in patients who are undergoing pharmacologic treatment for bipolar disorder.

**Significance Statement:** This study aims to find out whether suicide events that occur during the pharmacological treatment of bipolar disorder, a severe psychiatric disorder that is highly associated with suicide behavior, can be predicted. Using existing methods, we developed and compared several predictive models. We showed that these models performed similarly to predictive models of other outcomes, such as treatment efficacy, in unipolar and bipolar depression. This suggests that suicide events during bipolar disorder may be a feasible target for individualized interventions in the future.

## Introduction

Bipolar disorder is associated with elevated risks for suicide attempt and suicide death (Chen and Dilsaver), with a commonly cited lifetime risk for both at 15-25% (Oquendo et al., 2010). Several lines of indirect evidence suggest that on a population level, lithium may be the preferred agent for preventing suicidal behavior compared to other mood stabilizers (Cipriani et al., 2005; Baldessarini et al., 2006), although not all studies agree (Lauterbach et al., 2008), and discontinuation of lithium, especially rapid discontinuation, may increase risk for suicidal behavior (Muller-Oerlinghausen et al., 1992; Baldessarini et al., 1996; Baldessarini et al., 1999). Relatively little is known about the effect on suicidal behavior of anticonvulsant mood stabilizers such as valproate (Goodwin) and lamotrigine relative to lithium (Bowden et al., 2003; Calabrese et al., 2003). A double blind, parallel-group active comparison study (Oquendo et al., 2011) was conducted to compare lithium and valproate and did not show a relative advantage for lithium in preventing suicide attempt or suicide event, defined as suicidal ideation with a plan and requiring clinical intervention. While the study may have lacked statistical power due to the modest sample size, an alternative and additional explanation is that there may exist heterogeneities in treatment outcome: lithium might work better for some patients, while valproate might work better for others. Testing for moderator-treatment interaction, however, is also limited by the small sample size.

We sought to approach this problem by modeling treatment response heterogeneity directly. A clearer understanding of treatment response heterogeneity may allow researchers to develop individualized treatment rules to tailor treatment on an individual level (Qian and Murphy, 2011), and such strategies have shown promise in other areas of medicine such as oncology (Friedman et al., 2015) and cardiology (Antman and Loscalzo, 2016). Similar work has been emerging recently in psychiatry for major depression (Chekroud et al., 2016) autism (Abraham et al., 2017), and suicidal risk stratification using non-pharmacological datasets (Goldstein et al., 1991). A variety of treatment by baseline covariate interactions have been reported in depression: serotonergic antidepressants were reported to be more effective in patients with more severe depression (Fournier et al.), and, in particular, were more effective compared with a noradrenergic antidepressant in treating suicidal thoughts and behavior as baseline suicidal ideation severity increased (Grunebaum et al., 2012). While prospective observational studies identified a number of significant single predictors in major mood disorders (Oquendo et al., 2004; Sokero et al., 2005; Galfalvy et al., 2006) for suicide attempts and death, multi-predictor individual-level models based on large, and presumably heterogeneous datasets have been less successful (Goldstein et al., 1991; Marangell et al., 2006). Recently, new models using machine learning techniques and larger observational datasets, such as the Army Study To Assess Risk & Resilience in Servicemembers (STARRS) dataset, allow for more accurate risk stratification for individual level prediction of suicide risk after hospitalization (Kessler et al., 2015) and through outpatient mental health visits (Kessler et al., 2017b). These predictive models have also been built for electronic medical records (Kessler et al., 2017a). Nevertheless, these approaches have not been applied to treatment datasets, and therefore we do not know whether the predictive information can allow for tailoring of treatment, especially because so few clinical trials specifically targeted changes in suicidal thoughts and behavior as the primary outcome.

In the present study, we applied existing methodological approaches by building multi-predictor, predictive survival models of suicide event using a treatment dataset containing data from individuals undergoing pharmacological treatment of bipolar disorder. We sought to build models with both treatment group assignment and patient characteristics (i.e. models incorporating covariate by treatment interactions) have better predictive performance compared to models with only patient characteristics. This addresses not just the impact of treatment group on outcome, but also whether there could be improvement of outcome when treatment choice is tailored to individuals’ characteristics. The goal of the analysis is to provide a foundation that future studies targeting emerging suicidal ideations and behavior can also be plausibly designed to be more risk adaptive (Tomlinson et al., 2020).

## Material and Methods

### Trial Methodology Summary

This study was based on a pragmatic, parallel-group 2.5-year double-blinded trial for patients with bipolar disorder and past suicide attempts (N=98) (Oquendo et al., 2011). All participants provided written informed consent as approved by the New York State Psychiatric Institute Institutional Review Board. The trial enrolled a broadly inclusive group of patients with bipolar disorder (bipolar I or II disorder or bipolar disorder not otherwise specified, DSM-IV criteria); the participants were in a depressive or mixed episode; had at least one past suicide attempt; and between 18 to 75 years of age. Patients were also required to have had no history of nonresponse to adequate dosages of either lithium or valproate in the past 2 years. They were randomized at the start of the trial to either lithium or valproate with dosages optimized by an un-blinded physician based on blood levels. The design closely simulated clinical practice such that adjunct and second line treatment options followed existing guidelines for treatment of affective episodes in bipolar disorder. For further details on trial methodology, see (Oquendo et al., 2011).

### Baseline characteristics

Baseline psychopathology, demographic and patient characteristics were assessed using the Structured Clinical Interview for DSM-IV Axis I and II Disorders (SCID) as well as other structured inventories. Although patients were followed through a 2.5-year period, in the present study we only examined baseline predictors in developing our predictive models, based on the rationale that while early treatment efficacy might correlate with a good outcome, a predictive model based on only baseline characteristics would be more clinically useful.

### Outcome Measures

In the original trial, the primary outcome measures were time to suicide completion, time to suicide attempt and time to suicide event. Because there were no completed suicides in either group, and nonfatal suicide attempts were also rare, we examined time to suicide event as the prediction outcome. Suicide event was defined as either a suicide attempt (a potentially self-injurious behavior with some intent to end one’s life), or an episode of suicidal ideation with a plan, as assessed using a standardized inventory (Scale for Suicide Ideation), that required a change in treatment, such as addition of a rescue medication or hospitalization. This outcome was one of the primary outcome measures reported in the original study (Oquendo et al., 2011). Outcome measures were considered to have been at least partially observed for 94 subjects, with some censoring; the other 4 subjects did not return for follow-up visits and were not included in the subsequent analysis. Our choice of suicide event as the primary outcome rests on this outcome being an action response measure, whereas suicide attempts may be suppressed during a pragmatic clinical trial, when clinicians immediately intervene if a suicide attempt may be imminent.

### Statistical Analysis

The main goal of our modeling effort is to predict the time to suicide event for an individual patient given the treatment group assignment and other baseline patient characteristics. We used a mixed domain-knowledge (using existing literature in the suicide literature) and data-driven (depending on only the data to provide information on predictors) approach using a broad set of baseline covariates as predictors, similar to the approach proposed in (Kessler et al., 2015; Kessler et al., 2017b). However, unlike those studies, instead of screening all or most of the covariates recorded during the trial for associations with the outcome of interest and then empirically conducting model selection, we surveyed the existing literature to identify predictor covariates that had a theoretical basis for possibly influencing future suicide risk in bipolar disorder.

For each covariate, anywhere between 0 and 17% values were missing. We employed multiple imputation using the MICE package to impute missing covariates (Ian R White et al., 2011). Since we had a time-to-event outcome, we used the Nelson-Aalen estimator for the cumulative hazard of the survival time (rather than log of survival time) in the imputation model (I. R. White and Royston, 2009) along with all other clinical predictors under consideration. To account for potential interactions between covariates, we used classification and regression tree (CART) imputation (Doove et al., 2014) allowing all predictor variables including treatment group, event status, and the Nelson-Aalen estimator for the cumulative hazard. Using this procedure, we created 5 imputed data sets for analysis.

We constructed the models in the following stages: first we used the APA Guideline for Evaluation of Suicidal Behavior (Fawcett et al., 2009) and listed all the predictors that were proposed in the guideline as a relevant risk predictor during clinical assessment; we then conducted a focused literature search and included proposed predictors that appeared in at least an additional individual research article as a single predictor of suicide risk for any psychiatric condition (citation see Supplementary Material); finally, we cross-matched the proposed predictors to the trial dataset and included those predictors that corresponded to one or more of the items in the baseline assessments. Supplementary material provides definition of the covariates, evidence for inclusion, and the relevant citations (Supplementary Table 1).

In the second stage, we performed marginal screening to estimate the relationship between the outcome and each predictor individually. This allowed for comparing the predictors in single predictor analysis. We used Cox proportional hazard models to estimate the effect of each covariate and the effect of treatment on the final outcome (Table 1A, *single predictor models*) to explore potential predictor variables that have P-values less than 0.05 without specific adjustments for multiple comparisons. For each of the models in this stage and all subsequent stages, we fit a model to each of the five imputed data sets and pooled the estimates using Rubin’s rules (Marshall et al., 2009). We then built models with three additional sets of predictors: treatment group assignment of lithium vs. valproate, the predictor listed, and the predictor by treatment interaction terms (Table 1 B, *interaction models*). To evaluate models from the prediction point of view, we used a method proposed by Smith et al. (Smith et al., 2014) to obtain the optimism-adjusted Harrell’s C-statistic as the measure of the predictive performance of the single predictor model. The C-statistic is a commonly used statistical metric of discrimination for individual-level prediction models. In binary prediction models, the C-statistic is the area under the curve (AUC) for a Receiver Operating Characteristic (ROC), and it varies from 0.5 (prediction by chance only) to 1 (perfect accuracy). Harrell’s C is a generalization of this concept for survival data, that also varies from 0.5 to 1, and can be thought of as the average AUC for a time-dependent ROC curve, for classifying survival in time.

**Table 1.**
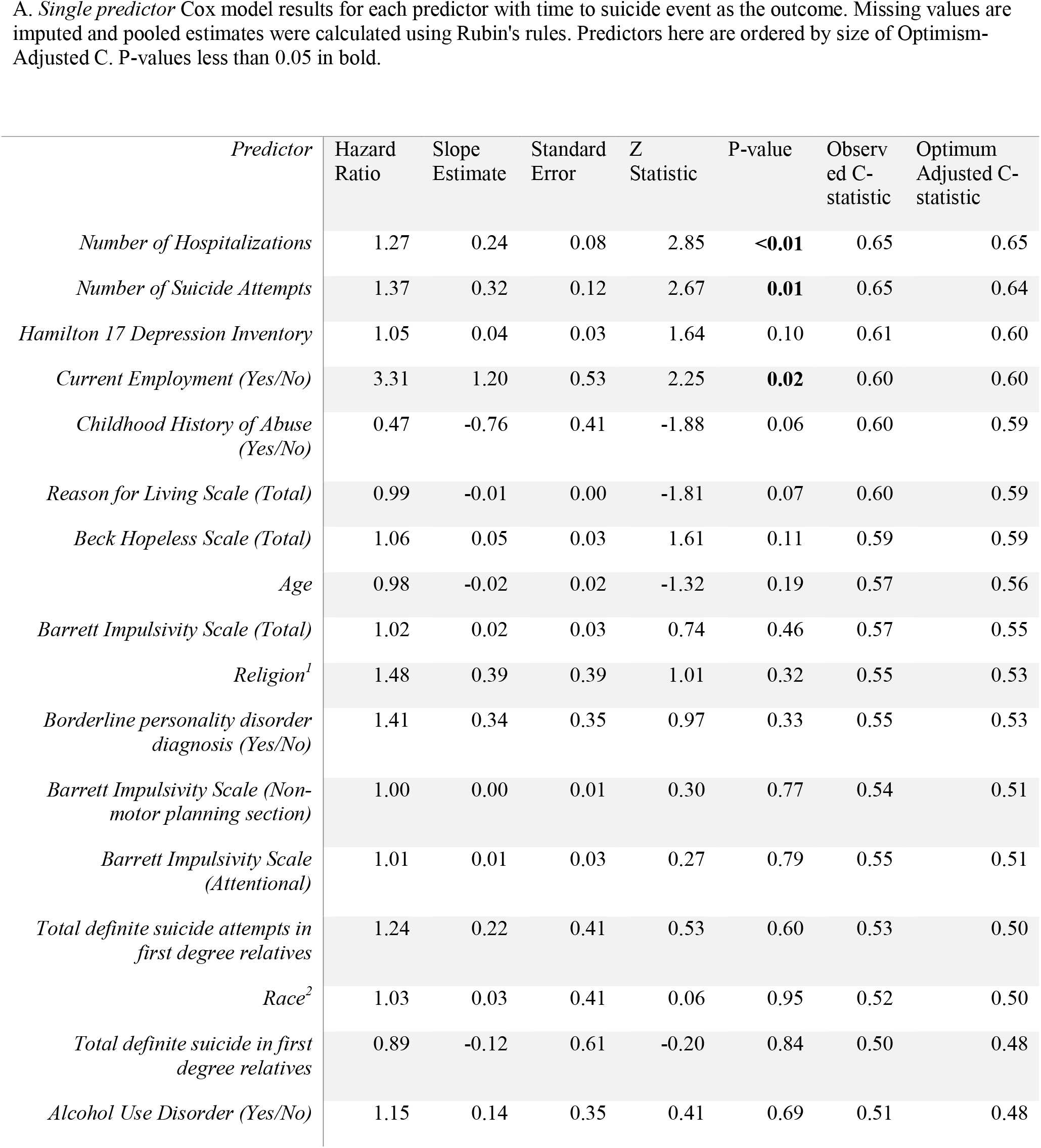
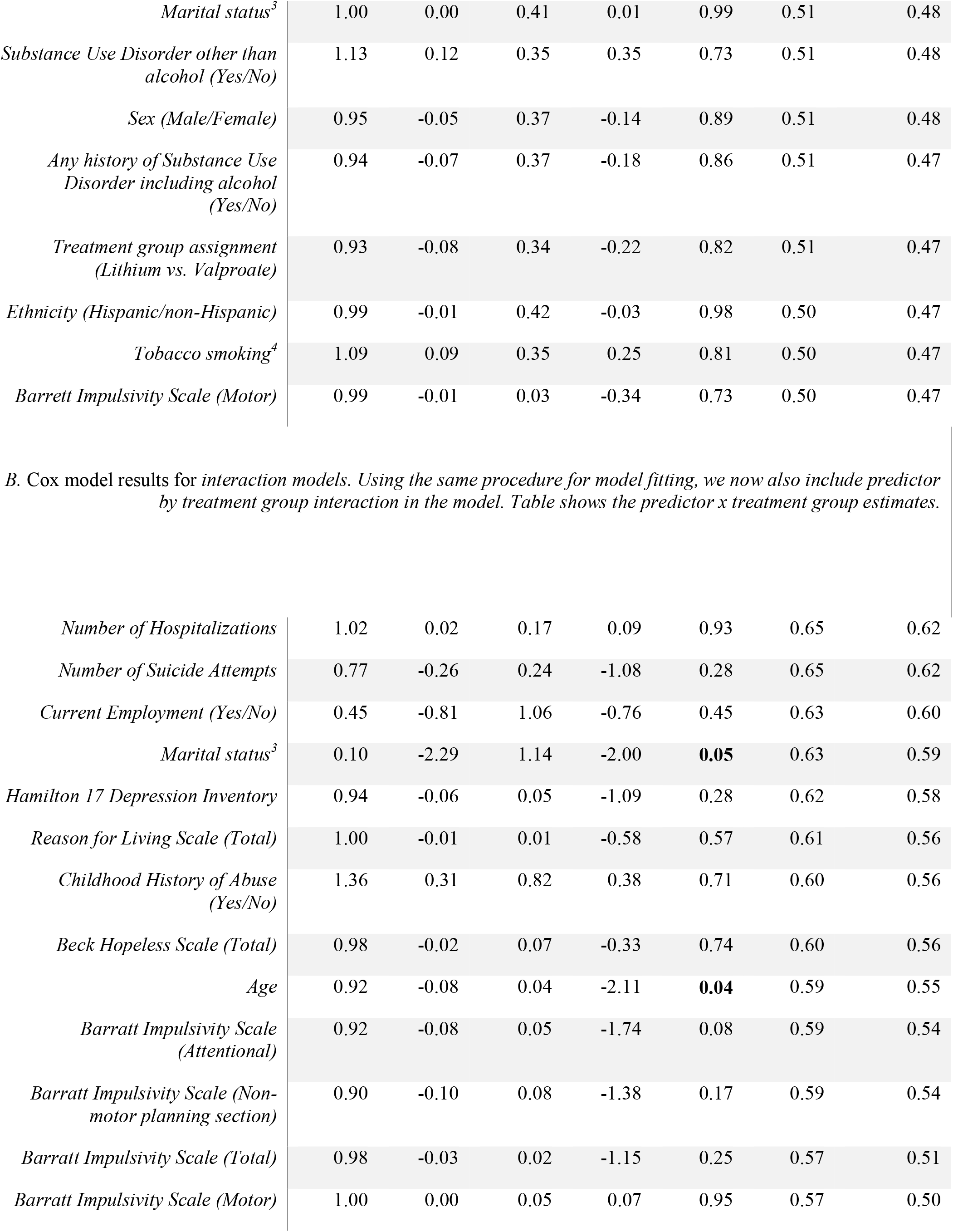
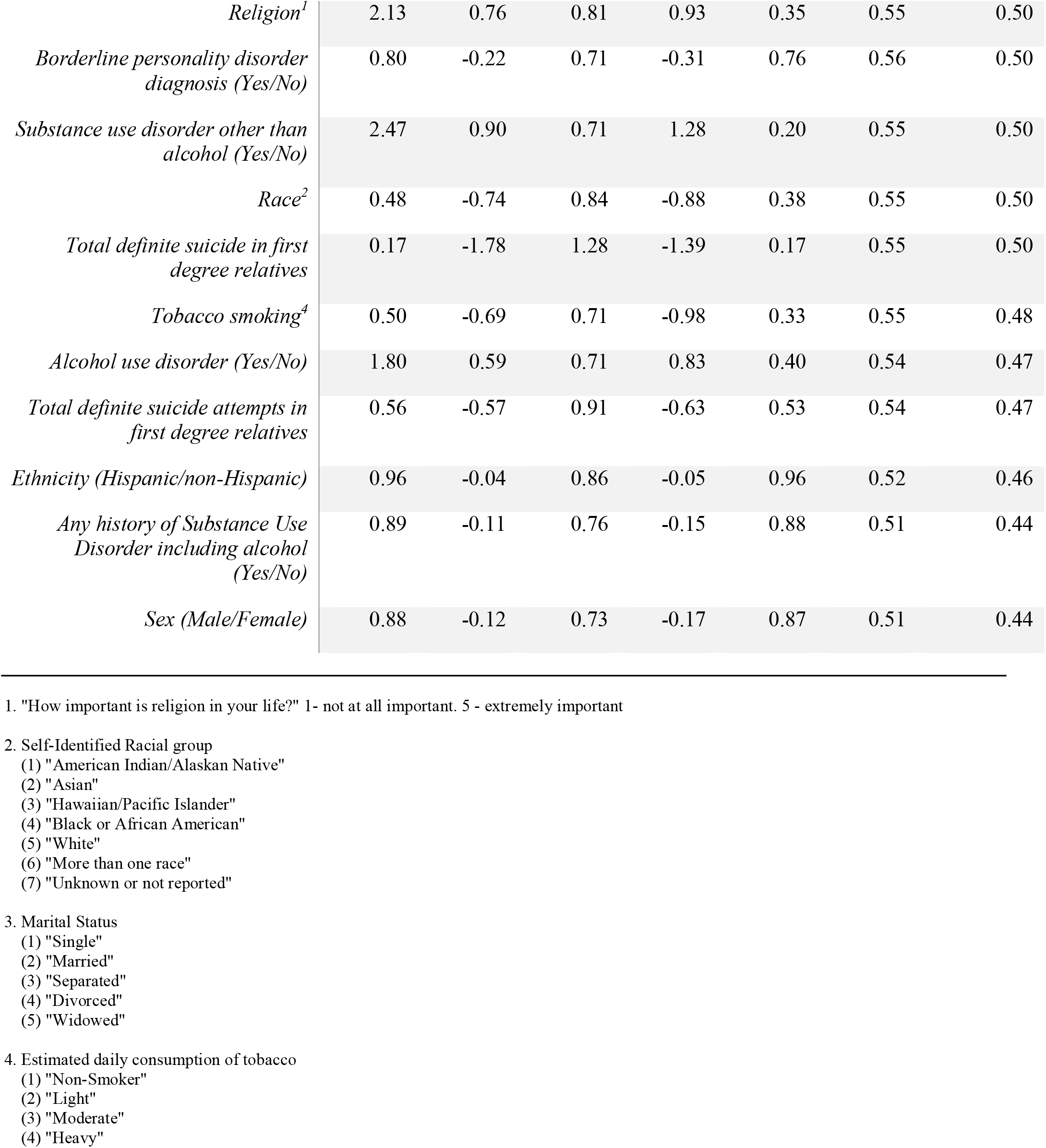
Marginal Screening for Predictively Informative Covariates. In the first stage, we selected variables that were previously described in the literature that potentially contain predictive information. We then created Cox Proportional Hazards *single predictor models* (A) for each predictor and *interaction models* (B) which contain the predictor, treatment group assignment, and the interaction of predictor and treatment group assignment.

| *Predictor* | Hazard Ratio | Slope Estimate | Standard Error | Z Statistic | P-value | Observed C-statistic | Optimum Adjusted C-statistic |
| --- | --- | --- | --- | --- | --- | --- | --- |
| *Number of Hospitalizations* | 1.27 | 0.24 | 0.08 | 2.85 | **<0.01** | 0.65 | 0.65 |
| *Number of Suicide Attempts* | 1.37 | 0.32 | 0.12 | 2.67 | **0.01** | 0.65 | 0.64 |
| *Hamilton 17 Depression Inventory* | 1.05 | 0.04 | 0.03 | 1.64 | 0.10 | 0.61 | 0.60 |
| *Current Employment (Yes/No)* | 3.31 | 1.20 | 0.53 | 2.25 | **0.02** | 0.60 | 0.60 |
| *Childhood History of Abuse (Yes/No)* | 0.47 | -0.76 | 0.41 | -1.88 | 0.06 | 0.60 | 0.59 |
| *Reason for Living Scale (Total)* | 0.99 | -0.01 | 0.00 | -1.81 | 0.07 | 0.60 | 0.59 |
| *Beck Hopeless Scale (Total)* | 1.06 | 0.05 | 0.03 | 1.61 | 0.11 | 0.59 | 0.59 |
| *Age* | 0.98 | -0.02 | 0.02 | -1.32 | 0.19 | 0.57 | 0.56 |
| *Barrett Impulsivity Scale (Total)* | 1.02 | 0.02 | 0.03 | 0.74 | 0.46 | 0.57 | 0.55 |
| *Religion<sup>1</sup>* | 1.48 | 0.39 | 0.39 | 1.01 | 0.32 | 0.55 | 0.53 |
| *Borderline personality disorder diagnosis (Yes/No)* | 1.41 | 0.34 | 0.35 | 0.97 | 0.33 | 0.55 | 0.53 |
| *Barrett Impulsivity Scale (Non-motor planning section)* | 1.00 | 0.00 | 0.01 | 0.30 | 0.77 | 0.54 | 0.51 |
| *Barrett Impulsivity Scale (Attentional)* | 1.01 | 0.01 | 0.03 | 0.27 | 0.79 | 0.55 | 0.51 |
| *Total definite suicide attempts in first degree relatives* | 1.24 | 0.22 | 0.41 | 0.53 | 0.60 | 0.53 | 0.50 |
| *Race<sup>2</sup>* | 1.03 | 0.03 | 0.41 | 0.06 | 0.95 | 0.52 | 0.50 |
| *Total definite suicide in first degree relatives* | 0.89 | -0.12 | 0.61 | -0.20 | 0.84 | 0.50 | 0.48 |
| *Alcohol Use Disorder (Yes/No)* | 1.15 | 0.14 | 0.35 | 0.41 | 0.69 | 0.51 | 0.48 |
| *Marital status*<sup>3</sup> | 1.00 | 0.00 | 0.41 | 0.01 | 0.99 | 0.51 | 0.48 |
| *Substance Use Disorder other than alcohol (Yes/No)* | 1.13 | 0.12 | 0.35 | 0.35 | 0.73 | 0.51 | 0.48 |
| *Sex (Male/Female)* | 0.95 | -0.05 | 0.37 | -0.14 | 0.89 | 0.51 | 0.48 |
| *Any history of Substance Use Disorder including alcohol (Yes/No)* | 0.94 | -0.07 | 0.37 | -0.18 | 0.86 | 0.51 | 0.47 |
| *Treatment group assignment (Lithium vs. Valproate)* | 0.93 | -0.08 | 0.34 | -0.22 | 0.82 | 0.51 | 0.47 |
| *Ethnicity (Hispanic/non-Hispanic)* | 0.99 | -0.01 | 0.42 | -0.03 | 0.98 | 0.50 | 0.47 |
| *Tobacco smoking*<sup>4</sup> | 1.09 | 0.09 | 0.35 | 0.25 | 0.81 | 0.50 | 0.47 |
| *Barrett Impulsivity Scale (Motor)* | 0.99 | -0.01 | 0.03 | -0.34 | 0.73 | 0.50 | 0.47 |
| *Number of Hospitalizations* | 1.02 | 0.02 | 0.17 | 0.09 | 0.93 | 0.65 | 0.62 |
| *Number of Suicide Attempts* | 0.77 | -0.26 | 0.24 | -1.08 | 0.28 | 0.65 | 0.62 |
| *Current Employment (Yes/No)* | 0.45 | -0.81 | 1.06 | -0.76 | 0.45 | 0.63 | 0.60 |
| *Marital status*<sup>3</sup> | 0.10 | -2.29 | 1.14 | -2.00 | **0.05** | 0.63 | 0.59 |
| *Hamilton 17 Depression Inventory* | 0.94 | -0.06 | 0.05 | -1.09 | 0.28 | 0.62 | 0.58 |
| *Reason for Living Scale (Total)* | 1.00 | -0.01 | 0.01 | -0.58 | 0.57 | 0.61 | 0.56 |
| *Childhood History of Abuse (Yes/No)* | 1.36 | 0.31 | 0.82 | 0.38 | 0.71 | 0.60 | 0.56 |
| *Beck Hopeless Scale (Total)* | 0.98 | -0.02 | 0.07 | -0.33 | 0.74 | 0.60 | 0.56 |
| *Age* | 0.92 | -0.08 | 0.04 | -2.11 | **0.04** | 0.59 | 0.55 |
| *Barratt Impulsivity Scale (Attentional)* | 0.92 | -0.08 | 0.05 | -1.74 | 0.08 | 0.59 | 0.54 |
| *Barratt Impulsivity Scale (Non-motor planning section)* | 0.90 | -0.10 | 0.08 | -1.38 | 0.17 | 0.59 | 0.54 |
| *Barratt Impulsivity Scale (Total)* | 0.98 | -0.03 | 0.02 | -1.15 | 0.25 | 0.57 | 0.51 |
| *Barratt Impulsivity Scale (Motor)* | 1.00 | 0.00 | 0.05 | 0.07 | 0.95 | 0.57 | 0.50 |
| *Religion<sup>1</sup>* | 2.13 | 0.76 | 0.81 | 0.93 | 0.35 | 0.55 | 0.50 |
| *Borderline personality disorder diagnosis (Yes/No)* | 0.80 | -0.22 | 0.71 | -0.31 | 0.76 | 0.56 | 0.50 |
| *Substance use disorder other than alcohol (Yes/No)* | 2.47 | 0.90 | 0.71 | 1.28 | 0.20 | 0.55 | 0.50 |
| *Race<sup>2</sup>* | 0.48 | -0.74 | 0.84 | -0.88 | 0.38 | 0.55 | 0.50 |
| *Total definite suicide in first degree relatives* | 0.17 | -1.78 | 1.28 | -1.39 | 0.17 | 0.55 | 0.50 |
| *Tobacco smoking<sup>4</sup>* | 0.50 | -0.69 | 0.71 | -0.98 | 0.33 | 0.55 | 0.48 |
| *Alcohol use disorder (Yes/No)* | 1.80 | 0.59 | 0.71 | 0.83 | 0.40 | 0.54 | 0.47 |
| *Total definite suicide attempts in first degree relatives* | 0.56 | -0.57 | 0.91 | -0.63 | 0.53 | 0.54 | 0.47 |
| *Ethnicity (Hispanic/non-Hispanic)* | 0.96 | -0.04 | 0.86 | -0.05 | 0.96 | 0.52 | 0.46 |
| *Any history of Substance Use Disorder including alcohol (Yes/No)* | 0.89 | -0.11 | 0.76 | -0.15 | 0.88 | 0.51 | 0.44 |
| *Sex (Male/Female)* | 0.88 | -0.12 | 0.73 | -0.17 | 0.87 | 0.51 | 0.44 |

1. "How important is religion in your life?" 1- not at all important. 5 - extremely important
2. Self-Identified Racial group
- (1) "American Indian/Alaskan Native" - (2) "Asian" - (3) "Hawaiian/Pacific Islander" - (4) "Black or African American" - (5) "White" - (6) "More than one race" - (7) "Unknown or not reported"
3. Marital Status
- (1) "Single" - (2) "Married" - (3) "Separated" - (4) "Divorced" - (5) "Widowed"
4. Estimated daily consumption of tobacco
- (1) "Non-Smoker" - (2) "Light" - (3) "Moderate" - (4) "Heavy"

In the final stage, we estimated the performance of different multi-predictor models for time to suicide event on each of the 5 imputed data sets and again used optimism-adjusted C-statistic as a performance measure:

MODEL 1: Cox model with main effects of predictors only. In this model we selected only those predictors with optimism-adjusted C-values ≥ 0.6 in the *single predictor models*.

MODEL 2: Cox model with main effect and predictor by treatment interactions with optimism-adjusted C-values ≥ 0.6 in either the *single predictor or interaction models*.

MODEL 3: We used LASSO with tuning parameters selected via 10-fold Cross Validation as an data-adaptive method for variable selection of all covariates with only main effect (Tibshirani, 1997). LASSO is a well-known algorithm that exploits a procedure known as L-1 minimization that penalizes the size of the coefficients in a regression model, and therefore typically selects for variables that have relatively strong predictive ability.

MODEL 4: We used LASSO fit as described for Model 3, but for variable selection for both main effect and predictor by treatment interaction terms.

MODEL 5: We employed a Survival Trees strategy (Taylor, 2011). Survival trees are a popular alternative to parametric and semiparametric models. They offer substantial flexibility in the construction of predictive models that allow the data, instead of the modeler, to identify relevant interactions between any of the variables being considered. In constructing survival trees, variable selection is automatic, and the fitted tree can be used to group subjects according to their survival behavior based on relevant covariates (Bou-Hamad et al., 2011). There is no obvious or agreed upon way to pool survival trees from multiply imputed data sets, and a general principle is to look for similarities on multiple trees to obtain a “consistent tree”. To demonstrate this, we plotted a Survival Tree with the corresponding number of events over time in each node (Fig. 1). We defined Variable Importance of a predictor as the mean rank for a predictor in all the Survival Trees obtained in imputed data sets (larger is better), and then ranked the variables from most important on average to least important. We then calculated the average C-statistic from all imputed datasets.

**Figure 1.**
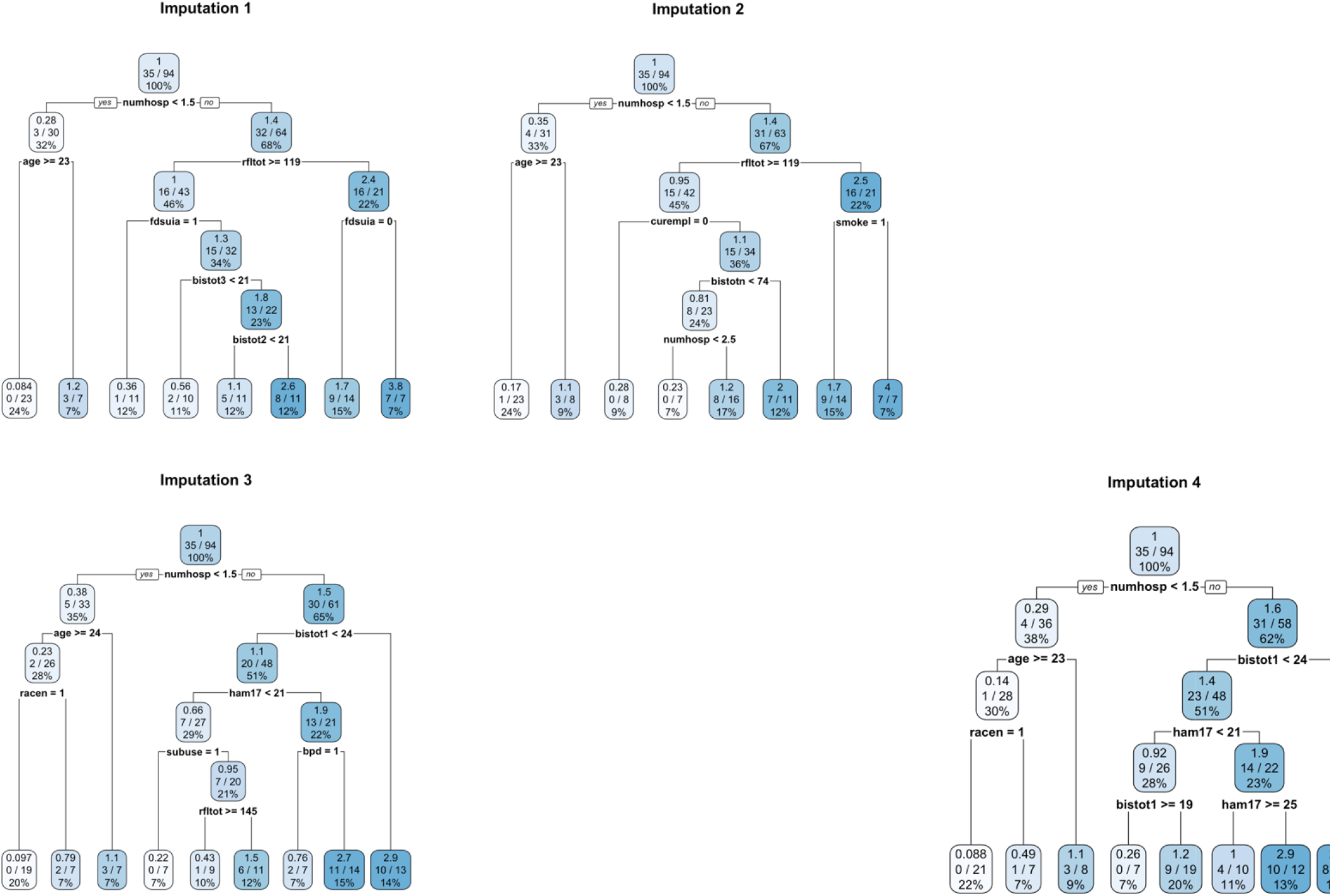
Survival Tree Analyses. Illustrative Survival Trees delineating various predictor variables as nodes (numhosp – Number of Hospitalizations, rfltot – Reason For Living (Total), fdsuia - Total definite suicide attempts in first degree relatives, bisttot1 – Barratt Impulsivity Scale (attentional), bisttot2 – Barratt Impulsivity Scale (Motor), bisttot3 - Barratt Impulsivity Scale (non-planning), bistoton—Barratt Impulsivity Scale (Total), curempl – Current Employment (1-Yes/0-No), smoke – Tobacco smoking (1-Yes/0-No)) and classification of differential risk groups. Top number in each box is the number of suicide events/time in the node; middle fraction shows number of suicide events out of number of subjects in the node; third number gives the % of participants from the whole sample in that node who had a suicide event. Each tree results in one imputation. Color of the bars indicate the proportion of participants who were partitioned into the branch, with darker color indicating a higher proportion.

All analyses were carried out using R Statistical Package (R version 2.12.1 Copyright (C) 2010 The R Foundation for Statistical Computing).

## Results

### Variable Selection and Marginal Screening

Using the methods outlined, we screened 24 predictors using the *single predictor (main effect only)* and *interaction with treatment group* models (Table 1A/B). We found that four factors reached an arbitrary threshold of optimism-adjusted C of 0.6 (number of hospitalizations, number of suicide attempts, 17-item Hamilton Depression Rating Scale (Ham17), Current Employment (Yes/No)). Number of prior hospitalizations, number of past suicide attempts and current employment status reached the typical p-value threshold of 0.05 based on their corresponding single predictor models. On the other hand, marital status and age appeared to show an interaction effect detected through a conventional p-value threshold of 0.05, but interaction model of neither predictor yielded an optimism adjusted C-statistic of 0.6 or greater, suggesting that the effect sizes of these interactions are insufficient for prediction. While both number of hospitalization and number of suicide attempts reached statistical significance in the *single predictor models*, the effect sizes (Hazard Ratio) were relatively modest (1.27 and 1.37 respectively).

### Predictive Model Performance

In model 1, we included the four predictors identified in the marginal screening stage. This multi-predictor Cox model yielded an apparent C of 0.697 and an optimism-adjusted C of 0.671, while 95% bootstrap confidence intervals (CIs) in each of the 5 imputed data sets showed that the smallest lower bound across those CIs was 0.571 and the largest upper bound was 0.800. Adding additional interaction terms (model 2) to this multi-predictor model did not enhance the predictive performance (optimism-adjusted C = 0.653, 95% CI smallest lower bound of 0.579 and largest upper bound of 0.795). In model 3, we used LASSO as a strategy to conduct automatic variable selection of all 24 predictors and obtained a model with an apparent C of 0.719 and an optimism-adjusted C of 0.62. Inclusion of the interaction terms did not appear to improve the performance of the LASSO based models. All of the predictors that appeared in the marginal screening strategy also were selected by the LASSO models.

### Survival Tree Modeling

We examined characteristics of the individual trees from each imputed dataset to look for similarities. In order to do so, we plotted survival trees derived from four imputed data sets (Fig. 1) for demonstration purposes. For each tree, we also computed the Variable Importance using recursive partitioning for each variable (Therneau et al., 2015), a classification process that repeatedly splits the sample into sub-samples based on dichotomous independent variables. Based on these importance measures, we assigned a rank to each variable and computed the mean rank for each variable across the 5 imputed data sets. Table 3 shows the top 10 variables in order of importance from most to least important corresponding to their average rank in the 5 imputed datasets (larger rank more important), providing an indicator of the most important predictors selected by the models. As with the other models, we computed the apparent and optimism-adjusted C value for each survival tree (one for each imputed data set) then took the average. The average apparent C-value is 0.815 and the optimism-adjusted C-value is 0.690.

**Table 2.**
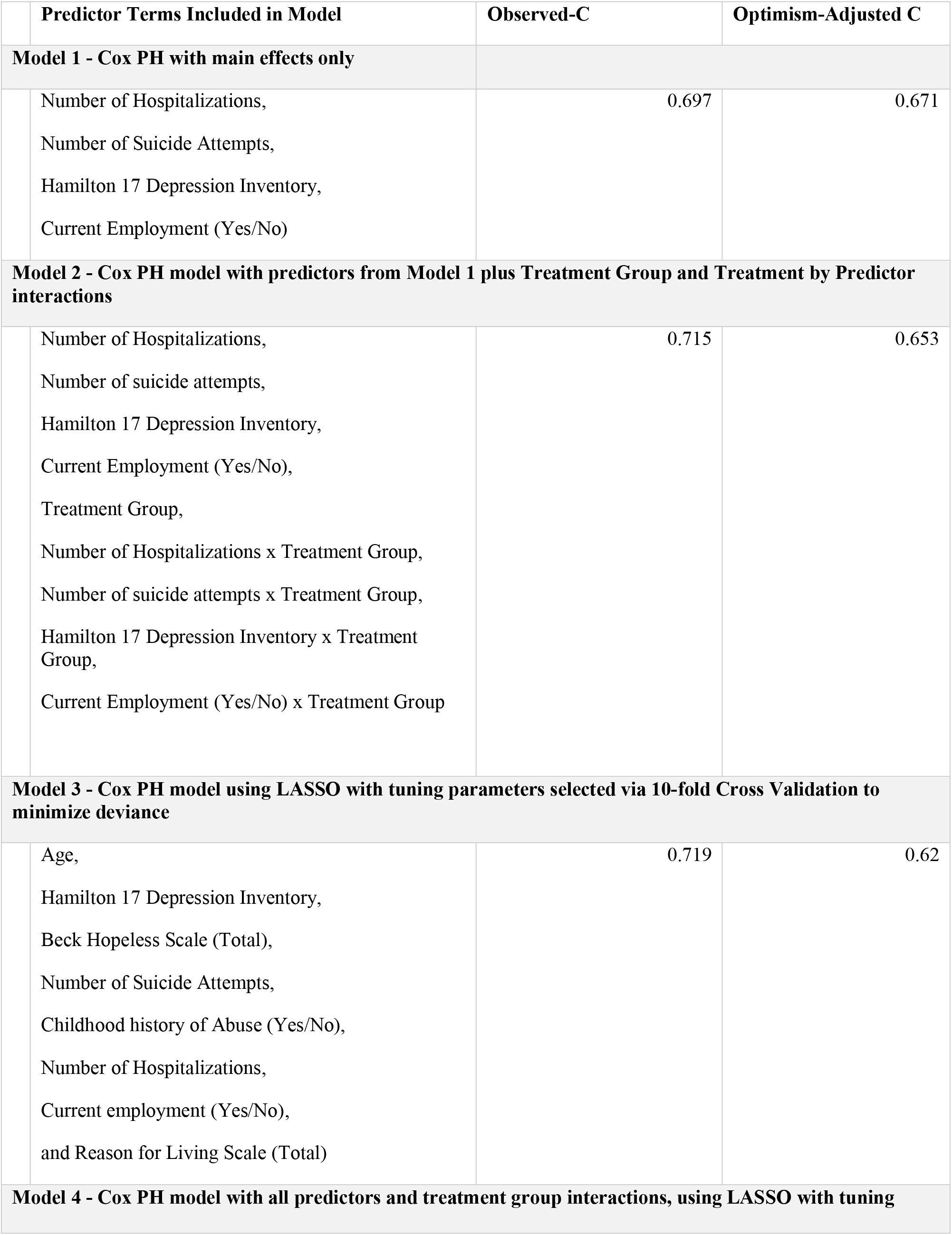
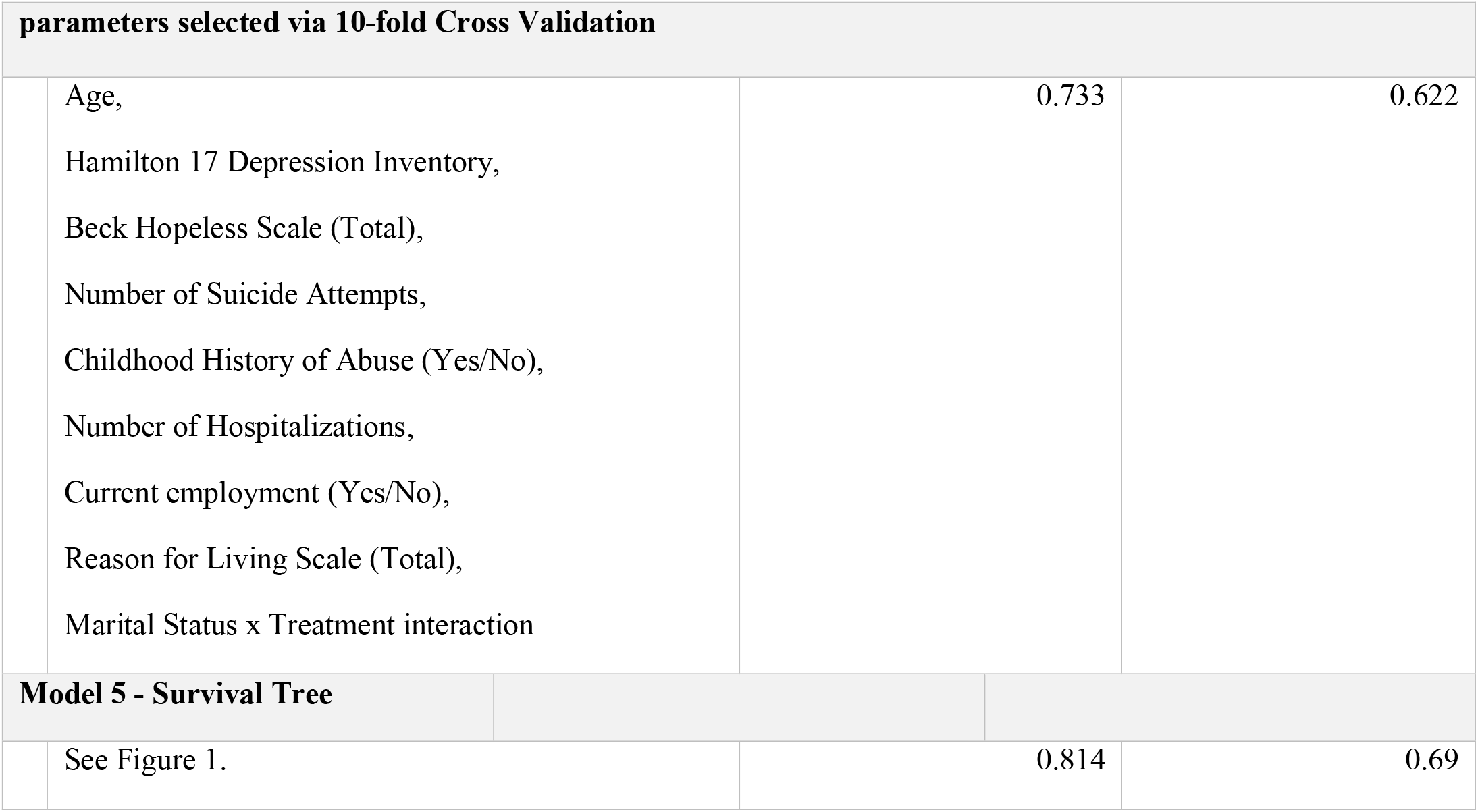
Summary Performance Measures of Predictive Models.

**Table 3.**
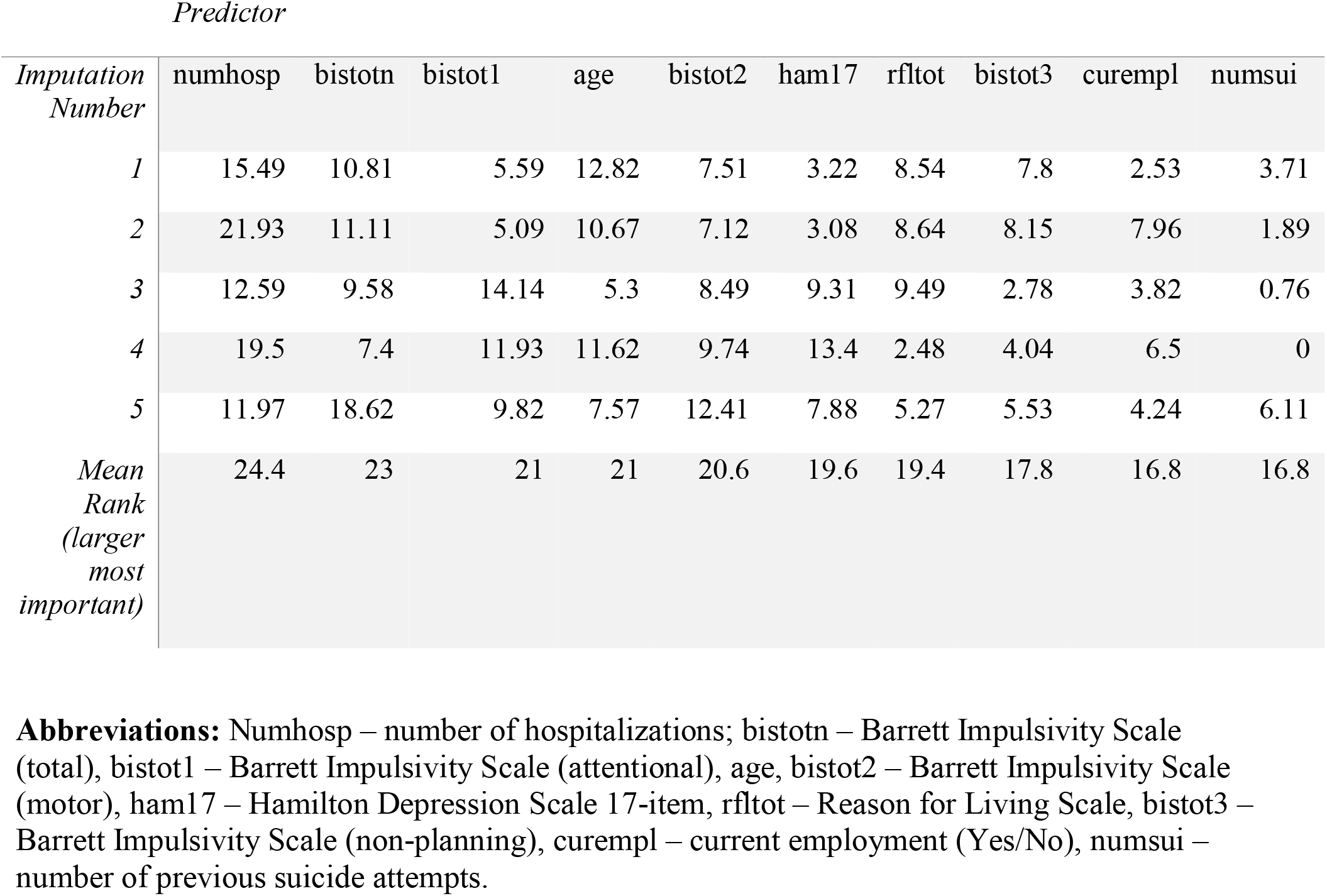
Variable Importance for the top variables most frequently appearing in the Survival Tree models (larger number is more important), ranked by the order they appear for each imputed dataset from which they were trained (larger number bottom row more important for a particular imputation).

| *Predictor* |  |  |  |  |  |  |  |  |  |  |
| --- | --- | --- | --- | --- | --- | --- | --- | --- | --- | --- |
| *Imputation Number* | numhosp | bistotn | bistot1 | age | bistot2 | ham17 | rfltot | bistot3 | curempl | numsui |
| 1 | 15.49 | 10.81 | 5.59 | 12.82 | 7.51 | 3.22 | 8.54 | 7.8 | 2.53 | 3.71 |
| 2 | 21.93 | 11.11 | 5.09 | 10.67 | 7.12 | 3.08 | 8.64 | 8.15 | 7.96 | 1.89 |
| 3 | 12.59 | 9.58 | 14.14 | 5.3 | 8.49 | 9.31 | 9.49 | 2.78 | 3.82 | 0.76 |
| 4 | 19.5 | 7.4 | 11.93 | 11.62 | 9.74 | 13.4 | 2.48 | 4.04 | 6.5 | 0 |
| 5 | 11.97 | 18.62 | 9.82 | 7.57 | 12.41 | 7.88 | 5.27 | 5.53 | 4.24 | 6.11 |
| *Mean Rank (larger most important)* | 24.4 | 23 | 21 | 21 | 20.6 | 19.6 | 19.4 | 17.8 | 16.8 | 16.8 |

**Abbreviations:** Numhosp – number of hospitalizations; bistotn – Barrett Impulsivity Scale (total), bistot1 – Barrett Impulsivity Scale (attentional), age, bistot2 – Barrett Impulsivity Scale (motor), ham17 – Hamilton Depression Scale 17-item, rfltot – Reason for Living Scale, bistot3 – Barrett Impulsivity Scale (non-planning), curempl – current employment (Yes/No), numsui – number of previous suicide attempts.

## Discussion

In this study, we built predictive models leveraging data from a randomized controlled trial of lithium versus valproate in treating patients with bipolar disorder who had a high propensity for suicidal behavior (Oquendo et al., 2011). In the original intention-to-treat analysis, no difference in treatment efficacy between lithium and valproate was detected with respect to time to suicide event. Using a predictive modeling approach, our analysis revealed two findings. First, we identified some clinically relevant characteristics that were predictors of outcome. In particular, we found that number of hospitalizations and number of previous suicide attempts, both commonly queried variables in clinical practice, were correlated with time to suicide events. We detected a significant predictor-treatment interaction for marital status and age, though incorporating these variables did not improve predictive performance. Second, we also showed we can predict suicide event with an optimism-adjusted C-value of 0.671 using 4 predictors in a Cox proportional hazards model. The use of more complex modeling, including treatment by baseline interactions, did not demonstrate significant improvement in predictive performance. These results have several implications, both methodologically and clinically. Previously, predictive modeling work for suicidal behavior relied primarily on observational datasets. Our results suggest that similar modeling strategies can be used for clinical trial datasets for bipolar disorder. The performance of our model is similar to work (Chekroud et al., 2016) in unipolar depression treatment response prediction, with similar individual level predictive performance: cross-validated C-statistic between 0.5 to 0.65 for various medications. A recent meta-analysis suggests that prediction models for bipolar and unipolar depression treatment response using clinical phenomenological predictors typically achieve a c-statistic between 0.65 and 0.75 (Lee et al., 2018). This suggests that while suicide events are in general a rare outcome, in enriched samples such as the one presented here, it may be feasible to use individual level predictive modeling as a technique to infer risk at the baseline, and design adaptive treatment trials specifically targeting suicidal behavior as an outcome.

Our results demonstrated that marginal variable selection, i.e. a data-driven approach based on single predictor models, can perform as well as or better than more complex variable selection methods, such as using LASSO, for sample sizes typical in suicide intervention studies. This is a consistent finding in application of machine learning to clinical trial datasets in other areas of psychiatry such as PTSD (Galatzer-Levy et al., 2014) and addiction (Luo et al., 2015). These examples of psychiatry treatment research, especially the single center studies, appear to lack sufficient sample size to constrain predictive models with large numbers of predictors. Failure to detect substantial improvement using complex non-parametric or non-linear modeling is also a common finding. For example, random forest models using 47 predictors and a simplified model using 15 predictors performed similarly (75% vs. 70%) in a study of prediction of treatment resistant depression (Kautzky et al., 2018). On the other hand, some datasets, such as those obtained from neuroimaging and genetics, may especially benefit from more complex feature modeling (Lee et al., 2018), and combining data sources may improve predictive performance (see recent reports from the EMBARC study (Trivedi et al., 2016; Petkova et al., 2017; Webb et al., 2018)).

Our results also raise several issues specific to predictive modeling for suicidal behavior. A recent meta-analysis reports that despite many years of research, individual risk factors do not provide enough information on their own to predict suicide, and prediction of suicide events over a long time period is disappointingly ineffective (Franklin et al., 2017). Several proposed solutions, including a more comprehensive survey of individual level characteristics through analyses of medical records, have yielded some early promising findings: a multi-stage approach for example, can detect future suicide attempts in U.S. Army soldiers who had no previous known suicidal ideation (Bernecker et al., 2018). Studies that involve electronic medical records often reach higher C-statistics (Kessler et al., 2017a), and these models are particularly useful in a large system in leveraging numerous alerts be sent to clinicians at lower cost for a heterogenous population. Our results suggest an additional approach: we may wish to identify those who are already diagnosed or being treated with a psychiatric disorder that make a suicide event much more likely. These enriched samples might allow us to treat these patients differently based on risk stratification. This is a secondary prevention strategy that is common in other fields of medicine, with corresponding risk prediction models and scores: e.g. Thrombolysis In Myocardial Infarction (TIMI) risk score for prediction of ischemic events for individuals who already have unstable angina; and the CHADS2 score (congestive heart failure, hypertension, age ≥75 years, diabetes mellitus, stroke [double weight]) for prediction of stroke for individuals who already have atrial fibrillation. For suicide outcomes, we believe the next step is to similarly construct more definitive predictive scores using larger, multi-center effectiveness trial datasets for similar high-risk psychiatric conditions (i.e. schizophrenia, bipolar disorder and borderline personality disorder). Secondly, an interesting unresolved question is whether results (and simple risk scores) from randomized trials generalize to observational studies of a much larger size, especially as many individuals followed in observational studies are treated with psychopharmacological agents—often they are not (Blanco et al., 2008). Thirdly, the potential to predict outcome and personalize treatment for patients with suicidal bipolar disorder may improve future trial design: we may therefore consider evaluating a specific suicide-targeted treatment program for individuals who are at high risk of suicide event as predicted by these models.

There are a number of limitations of this study. First, the study included only those bipolar patients with a prior suicide attempt, thus it may not generalize to the prediction of a first attempt. We deliberately limited our predictors to baseline characteristics, as they were the most applicable in clinical practice as a means to stratify patients at initial diagnosis. However, predictors that partially indicated treatment success, such as adherence records and early symptomatic resolutions, might also be useful predictors of eventual treatment outcome. Fitting and validating predictive models with a small dataset has a number of technical limitations (Braga-Neto and Dougherty, 2004; Isaksson et al., 2008), and may be prone to false positives and generate an overly confident estimate of predictive performance (Button et al., 2013)—this is partially addressed by using the optimism-adjusted C metric (Smith et al., 2014), though this method still underestimates true error rate, and more importantly, variance of the error rate. The promising conclusions of this study therefore would require external validation in a larger sample study. Finally, adoption of externally validated predictive models in clinical practice would involve centralized resources, such as the Memorial-Sloan Kettering Prediction Tools (Vickers, 2011), that have been widely used in other medical specialties but are not yet available in psychiatry.

## Supporting information

IRB APPROVAL NOTICE

ETHICS STATEMENT

## Data Availability

All data, methods and analysis scripts are available upon request.

## Acknowledgements

None

