## Supplementary Material for "Individual level prediction of emerging suicide events in the pharmacologic treatment of bipolar disorder"

Supplementary Section 1. Labels and explanations for predictors as well as summary citation for existing literature suggesting a possible association with suicidal behavior in observational studies.

Supplementary Table 1.

| AGE | While older adults have a higher completed suicide rates, younger adults are more likely to attempt suicide | [1] |
| --- | --- | --- |
| SEX (sex = 1 if male 0 if female) | In the US, suicide rates for men is four times as much as women. | [20] |
| SEX * AGE | Increasing suicide rates are especially pronounced for men as the age increases. | [3] |
| ETHNIC | In the US, the highest suicide rates occur in those over age 65 among non-Hispanic whites, Hispanics, and Asian-Pacific Islanders | [1] |
| RACEN | Race and ethnicity are variously defined as above in different studies |  |
| MARSTAT | **Marital status**: divorced, separated, or widowed individuals have rates four to five times higher than married individuals. | [4] |
| Ham21 | **21-item Hamilton depression score**: major mood disorder is strongly associated with increased risk of suicide. | [8] |
| Alcohol (1 – history of alcohol use disorder, 0-no history) | Suicide rates for patients with alcoholism are approximately six times those of the general population | [6] |
| Subuse (1 – history of any substance use disorder, 0 – no history) | Abuse of other substances is associated with increased rates of suicide. | [7] |
| Bpd (0 – not diagnosed, 1-diagnosed) | Patients with **borderline personality disorder** have a higher risk | [20] |
| bpd*alcohol | Comorbid substance abuse disorder may contribute to the overall risk | [9] |
| HOPETOTN | **Beck hopelessness scale:** hopelessness is a dimension that is associated with increased rates. | [10] |
| BISTOT1, BISTOT2, BISTOT3, BISTOTN | **Barratt Impulsiveness Scale in the 3 sections: attentional, motor, non-planning.** There is strong evidence for a role of impulsivity in self-injurious behavior. | [11] |
| NUMSUI (numerical) | Any past suicide attempt is one of the major risk factors for future suicide attempts. | [12] |
| ABUSE (1 – present of a history of childhood abuse, 0 – no such history) | Suicide rates are increased at least 10-fold in those with a history of childhood abuse. | [13] |
| ABUSCOD (1-childhood history of sexual abuse, 0 – no such history) | There maybe a specific increase of risk for sexual abuse. | [14] |
| ABUSEPL | The relationship of the abuser may also be a risk factor. | As above |
| NUMHOSP (numerical) | Number of hospitalization is predictive for a future hospitalization for suicide attempt. | [15] |
| FDSUI, FDSUIA | Family history of suicide is associated with an increased risk | [16] |
| CUREMPL (1-employed, 0 – not employed) | Unemployment is associated with an increase in rate. | [17] |
| RELIGION | Some evidence points to religion as protective. | [18] |
| RFLTOT | **Reason for living scale**: an ability to cite reasons for living is protective | [19] |
| ALCOHOL  INTERACTIONS | In patients with a history of alcohol dependence, there is evidence for greater rates in with female sex, younger age, lower economic status, unemployment, living alone, poor social support, other psychiatric disorders, personality disturbance, and other substance use | [20, 21] |

[2] Web-Based Injury Statistics Query and Reporting System, National Center for Injury Prevention and Control, Centers for Disease Control and Prevention: Fatal injury data for 2000.

[3] Institute of Medicine: Reducing Suicide: A National Imperative. Washington, DC, National Academies Press, 2002.

[10] Brown GK: A Review of Suicide Assessment Measures for Intervention Research With Adults and Older Adults. Rockville, Md, National Institute of Mental Health, 2002.
