## Supplementary material for "Individual level prediction of emerging suicide events in the pharmacologic treatment of bipolar disorder": IRB APPROVAL NOTICE

**Subject:** PRISM No Further Review Required for 8107 PSF-Individual-level prediction of emerging suicide events in the pharmacologic treatment of bipolar disorder_(12/11/2020-SL)
**Date:** Friday, December 18, 2020 at 3:52:21 PM Eastern Standard Time
**From:** PRISM for Corinne Rogers
**To:** Luo, Sean (NYSPI)

Dear Sean Luo,

Thank you for using PRISM. The following study does not require any further review.

Protocol #: **8107**
Title: **Individual-level prediction of emerging suicide events in the pharmacologic treatment of bipolar disorder**

Click here to view any correspondence attached to this protocol.

Thank you
NYSPI-IRB

Protocol Title:
**Individual-level prediction of emerging suicide events in the pharmacologic treatment of bipolar disorder**

Version Date:
**01/05/2021**

Protocol Number:
**8107**

First Approval:
**N/A**

Expiration Date:
**Not yet accepted**

Contact Principal Investigator:
**Sean Luo, MD, PHD**
****

Research Chief:
**J. John Mann, MD**

### Cover Sheet

Choose ONE option from the following that is applicable to your study
If you are creating a new protocol, select "I am submitting a new protocol." As 5 Year Renewals are no longer required, this option remains for historical purposes.
I am submitting a new protocol

### Division & Personnel

#### Division

What Area Group does the PI belong to?
What Division/Department does the PI belong to?
Substance Use Disorders
Within the division/department, what Center or group are you affiliated with, if any?
None

#### Unaffiliated Personnel

List investigators, if any, who will be participating in this protocol but are not affiliated with New York State Psychiatric Institute or Columbia University. Provide: Full Name, Degrees and Affiliation.
Not applicable

### Procedures

**To create the protocol summary form, first indicate if this research will include any of the following procedures**

✓ Secondary Data Analysis Only

Does the data contain Protected Health Information (PHI)?
No

### Research Support/Funding

Will an existing internal account be used to support the project?
No
Is the project externally funded or is external funding planned?
No

### Studies Involving Only Analysis of Previously Collected Information

Is interaction with human subjects or collection of data ongoing or planned at this time?
No
Is the data publicly available?
No
Was the data collected for research purposes?
Yes
*Was the data collected with IRB approval and informed consent?*
Yes
Does the data include unique identifiers or codes which link the data to subject identity?
No
*The research does not appear to be human subjects research. Please complete this and the Lay Summary page.*

### Lay Summary of Proposed Research

Lay Summary of Proposed Research
A previous study studied effect of using valproate vs. lithium as a treatment of bipolar disorder (N= 98, Oqeundo, 2011). This study is a follow-up study that used the individual level treatment data to identify predictors of treatment response, as well as follow up to explore whether a predictive model can estimate the probability of a suicide event.

### Persons designated to discuss and document consent

Select the names of persons designated to obtain consent/assent
Type in the name(s) not found in the above list

### Methods to Protect Confidentiality

Describe methods to protect confidentiality
No PHI is contained in the dataset being analyzed.
*Will the study be conducted under a certificate of confidentiality?*
No

### Uploads

Upload copy(ies) of the HIPAA form
Upload any additional documents that may be related to this study
