## Supplementary material for "Individual level prediction of emerging suicide events in the pharmacologic treatment of bipolar disorder": ETHICS STATEMENT

COLUMBIA PSYCHIATRY

JEFFREY A. LIEBERMAN, M.D.
LAWRENCE C. KOLB PROFESSOR AND CHAIRMAN
COLUMBIA UNIVERSITY DEPARTMENT OF PSYCHIATRY
DIRECTOR, NEW YORK STATE PSYCHIATRIC INSTITUTE

1051 RIVERSIDE DRIVE
NEW YORK, NY 10032

WWW.COLUMBIAPSYCHIATRY.ORG

Ethics Statement

MS ID#: MEDRXIV/2020/246603
MS TITLE: Individual level prediction of emerging suicide events in the pharmacologic treatment of bipolar disorder

This is a statement regarding the above manuscript.

1. Full names / affiliations of all ethics committees / Institutional Review Boards that ruled on ethics of your study.

This study is reviewed by the New York State Psychiatric Institute Institutional Review Board.

2. Decision made, i.e. whether ethical approval was given or waived.

Ethical approval was waived.
